# Therapeutic mammoplasty as a viable surgical approach in Breast Cancer Patients from India: A Single Institutional Audit

**DOI:** 10.1101/2021.06.22.21258390

**Authors:** C. B. Koppiker, Sneha Joshi, Rupa Mishra, Devaki A. Kelkar, Pragnya Chigurupati, Anjali Joshi, Jisha John, Shweta Kadu, Nutan Gangurde, Deepti Thakkar, Gautam Sharan, Upendra Dhar, HariKiran Allampati, Smeeta Nare, Ankush Dewle, Sanket Nagarkar, Laleh Busheri

**Affiliations:** Prashanti Cancer Care Mission, Pune, India; Centre for Translational Cancer Research: A Joint Initiative of Indian Institute of Science Education and Research (IISER) Pune and Prashanti Cancer Care Mission (PCCM) Pune, India; Orchids Breast Health Centre, a PCCM initiative, Pune, India; International School of Oncoplastic Surgery, Pune, India; Department of Radiation Oncology, Inlaks and Budhrani Hospital, Pune, India; Ruby Hall Clinic, Pune, India

**Author notes:** **Corresponding author** Chaitanyanand B. Koppiker M.D., Medical Director, Orchids Breast Health, Pune, India, Managing Trustee, Prashanti Cancer Care Mission, Pune, India, Research Lead, Centre for Translational Cancer Research (CTCR), Pune India, Honorary Associate Professor, University of East Anglia, UK, Director, Oncosciences, Jehangir Hospital, Pune, India, Founding Director, BreastGlobal Network, Board of Directors and Visiting Faculty, School of Oncoplastic Surgery, CA, USA, Member, Board of Directors, Indian Cancer Genome Atlas (ICGA), **Contact:**. These Authors have contributed equally (Sneha Joshi, Rupa Mishra, Devaki A. Kelkar). **Author Contributions:** CBK initiated the study concept and study design. PC, GS, UD, HA assisted with study design. AJ, NG, JJ, SN, DT, SJ, RM assisted with data acquisition. SJ, RM, DK, PC, CBK provided data analysis, interpretation while SJ, RM, DK, PC, LB assisted for quality control of data and algorithms. SK, DK, AJ, JJ, SJ, RM assisted in statistical analysis. SJ, RM, CBK wrote the manuscript while manuscript editing was done by SJ, RM, SN, AD. SJ, RM and DK provided critical manuscript review.

## Abstract

**Introduction:** Therapeutic mammoplasty (TM) is a type of oncoplastic breast surgery (OBS) well suited to breast cancers in medium-large sized breasts with ptosis, and in some cases of large or multifocal/Multicentric (MF/MC) tumors. It includes contralateral symmetrisation. This report describes our experiences and outcomes of TM in breast cancer patients in a single institutional cohort in India.

**Methods:** We present data for 207 cases (194 breast cancer, 13 benign disease) who underwent TM as part of their primary treatment. All patients underwent surgery after careful analysis of feasibility by a multidisciplanary tumor board and patient counselling. We report the clinicopathological profiles, surgical and oncological outcomes, and patient related outcome measures (PROMs) with different TM surgical procedures.

**Results:** Patients were relatively young at a median age of 49 years with moderate-large breasts and grade II-III ptosis. Patients underwent simple (n=96), complex (n=79) or extreme TM (n=46). Low post-operative complication rates and good-excellent cosmetic scores were observed. With median follow-up of 26 months, 148 patients completed more than 1 year follow-up. The 1-year BREAST-Q PROMs revealed good-to-excellent scores for all types of therapeutic mammoplasty.

**Conclusion:** We conclude that in a country where women present with large and locally advanced tumours, TM safely expands the indications for breast conservation surgery. PROMs scores show that this surgery is perceived to be physically and mentally satisfactory. With the popularization of this procedure, it is possible that more Indian patients with breast cancer will receive the benefits of breast conservation while maintaining their quality of life.

## Introduction

Breast conservation therapy (BCT) which involves breast conservation surgery (BCS) followed by adjuvant radiotherapy (RT) is now established as a standard of care for breast cancer^1,2^. Several studies have shown equivalent survival rates between BCS and mastectomy after a 20-year follow-up^3,4^. Recent studies have suggested a better disease free and overall survival with improvement in quality of life (QoL) in patients undergoing BCT as compared with mastectomy^5,6^.

However, unsatisfactory cosmetic outcomes post-BCT have been observed in cases where large excisions of the breast tissue were required.^7^. Such poor cosmesis that is a labelled as post-BCS breast defect or asymmetry between the breasts or nipples limits the application of conventional BCT. In addition, BCT has limited applications in patients with multi-focal or multi-centric (MF/MC) disease, and extensive microcalcifications. Conventionally, MF/MC cancers have been labelled as a contraindication for BCS, though low recurrence rates have been observed in such patients when treated with BCT^7^.

The concept of Oncoplastic Breast Surgery (OBS) was first introduced in the 1990’s by Prof. Audretsch when he described the technique of partial reconstruction of the breast, using plastic surgical techniques^8,9^. OBS is now increasingly being accepted as standard-of-care in surgical management of breast cancer cases across the world due to benefits such as oncological safety with concurrent improvement in aesthetic results and Quality of Life (QoL)^10–12^.

OBS procedures involving partial breast reconstruction are classified as volume replacement or volume displacement techniques^13,14^. Therapeutic Mammoplasty (TM) is a commonly used volume displacement technique suitable for OBS in women with medium/large breasts with ptosis. TM combines the advantages of oncologically safe wide excision of tumor with breast reduction, mastopexy and contralateral symmetrisation techniques^15^. TM has been shown to achieve satisfactory outcomes by reducing the breast size thereby facilitating better delivery and distribution of RT regimens, achieving contralateral breast symmetry, and improving the QoL. The oncological safety and efficacy of TM have been confirmed in early breast cancer cases indicated by higher rates of overall survival (OS), disease free survival (DFS) with low recurrence, lower complication rates and superior cosmesis^16^. Furthermore, TM offers an option for BCS in women who present with locally advanced breast cancer (LABCs) (>5cms), MF/MC or extensive microcalcifications wherein a mastectomy would be the surgical procedure of choice^15,17^. However, even though indicated for smaller ptotic breasts in selective cases, TM may not be effective due to paucity of the breast tissue^18^.

The Therapeutic Mammoplasty study (TeaM Study) was a large, international, multicentric cohort study to evaluate the safety and efficacy of TM in women undergoing BCS for invasive or pre-invasive breast cancer as per consensus guidelines developed by European breast units^17^. The TeaM study reported that the TM procedure is a safe and effective alternative to standard BCS or mastectomy. Recently, data from the national iBRA-2 and TEAM studies were combined to compare the safety and short-term outcomes of TM and mastectomy with or without immediate breast reconstruction (IBR). This data indicated that BCS was possible in 87% TM cases without delay in adjuvant treatment indicating that TM may allow high-risk patients who are not candidates for IBR to avoid mastectomy safely^17,19^.

Majority of Indian breast cancer patients present with large tumors in advanced stages^20^. This limits the scope of upfront BCS with or without OBS, unless the patient has a favorable breast-to-tumor ratio. In such patients, OBS with the TM procedure has been shown to effectively extend the boundaries of surgical excisions^21^. However, the field of OBS is still nascent in India and is practiced by a handful of breast surgeons in metropolitan cities.

With this background, we undertook the current study to investigate and analyze the outcomes of TM with a focus on oncological safety and efficacy. From our single-institutional TM cohort, we present data on 207 patients who underwent a TM after analysis of the feasibility and safety of the procedure and careful counselling and MDT discussion. In our practice, we classify TM techniques into 4 categories according to indications described in Table 1^13,14,22^.

**Table 1:** Types of Therapeutic Mammoplasty

| Type | Description | References |
| --- | --- | --- |
| Simple | For tumors within the reduction pattern (i.e., at 06'O clock)<br>The nipple is placed on the superior, supero-medial, and inferior pedicles, which are commonly used pedicles. | (Savalia and Silverstein 2016)<sup>18</sup> |
| Complex | For tumors outside the pattern of reduction (i.e., between 12 – 03 O' clock position (left breast) and 12 – 09 O clock (right breast)<br>Dual pedicle technique is applied in which extended and/or secondary pedicles (inferior, infero-lateral, or infero-medial) act as fillers which enhance the vascularity as an added advantage*.<br>*Extended or Secondary pedicles are the other parts of the breast that are generally excised, which are used to fill the defects. The later are preferred as they have better blood supply reaching the most distant areas of the pedicle as compared to extended ones | (Savalia and Silverstein 2016)<sup>18</sup> |
| Extreme | Include large multi-centric, multi-focal tumors, extensive DCIS and poor response to NACT requiring large areas of resection (>5cm), in which mastectomy would be the surgical procedure of choice. | (Silverstein et. al. 2015, Silverstein et al. 2016, Koppiker et al. 2019)<sup>21,29,31</sup> |
| Split Reduction | For tumors which lie outside the reduction pattern wherein the skin needs to be resected due to involvement or close proximity with tumor<br>The lower limb of the wise pattern is shifted over the tumor site. Then the outer limb of the wise pattern is shifted upwards to lie over the tumor so that there is no incision in the IMF on the outer side<br>In contrast to the other techniques, the incisions on the IMF and the horizontal limb on the side of the skin excision are omitted to preserve the vascularity and restructure the breast. | (Silverstein et al. 2015)<sup>30</sup> |

Based on the guidelines of the TeaM Study protocol, we report the clinicopathological profiles, outcomes and patient related outcome measures (PROMs) of our cohort and experiences related to various TM surgical techniques.

## Methods

### 1. Patient Selection

At our institution, routine pre-op counselling is performed by the surgeon to discuss options such as mastectomy, BCS and OBS. If the patient consents for TM, simultaneous contralateral breast reduction is also discussed This option gives superior cosmetic outcomes for women with ptotic breasts (Grades I-III) and moderate-to-large sized breasts.

### 2. Surgical Procedures

#### 2.1 Pre-operative Markings

In the pre-operative planning, appropriate markings are made on both breasts based on a wise pattern or vertical scar incision. The nipple areolar complex (NAC) is re-positioned between 19-23 cm from the sternal notch, which is often determined by placing the fingers at the inframammary fold (IMF) and projecting on the anterior surface of the breast into the meridian.

#### 2.2 Tumour Localization

Tumor localization is performed either pre-operatively by stereotactic guide-wire placement or by placement of wire and needle on the operating table using a high-resolution ultrasonography. In post-NACT cases, tumour is localized mid-NACT (at ∼ 1cm tumour size) by USG guided insertion of Liga clips (Koppiker et al., unpublished observations).

#### 2.3 Incision, Tumour Excision and Oncological Clearance

The surgery begins by marking out the wise pattern incision (Figure 1a). The wise pattern is located to excise the localized tumour with wide margins. The area of the appropriate pedicle which will carry the nipple is marked and de-epithelized. The tumor is then excised with wide margins through one of the limbs of the wise pattern. If required, further imaging of the specimen is performed using specimen mammography to ensure that the tumor is excised with wide margins The shaved margins of the cavity are further excised and sent for frozen sections to assess margin positivity. If any margins are close to the tumor (<0.5 cms), they are re-excised. Once negative tumor margins of the excision cavity are achieved the decision is made to use one of the appropriate pedicles.

**Figure 1:**
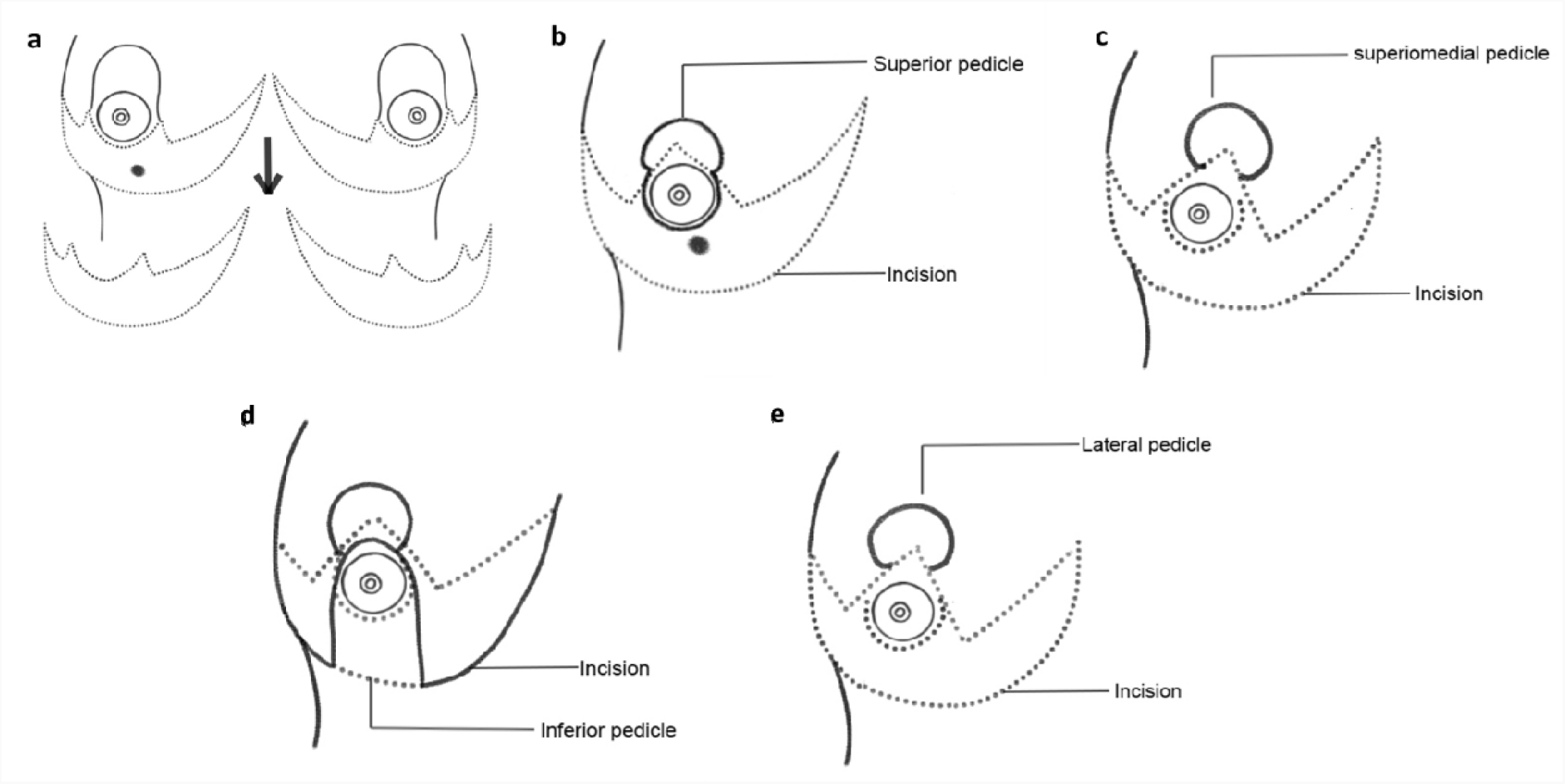
Schematic Representation of wise-pattern incision and various choices of pedicles. **a**. Wise-pattern incision **b**. superior pedicle **c**. superior-medial pedicle **d**. inferior pedicle **e**. lateral pedicle

#### 2.4 NAC Positioning

The NAC is marked out, and an incision is carefully made around the areola. The tumor and its quadrant are then widely excised through one of the limbs of wise pattern incision, (based on the type of TM technique decided).

#### 2.5 Marking Out the Tumor Bed for Targeting Radiotherapy

The tumor bed is marked with Liga clips in the superior, inferior, medial, and lateral, the basal and anterior margins. In our experience, the tumor margins remain contained within the initial tumor volume for targeted radiotherapy. The possibility of tumor margin getting repositioned in some other quadrant is less likely.

#### 2.6 Choice of Pedicles for Various TM Techniques

The appropriate pedicles are marked out and dissected according to the location of the tumor. According to quadrant diagrams (Fig. 1b-1e), if the tumor is at 12 O’clock position in UOQ, an extended inferior pedicle is used. If a tumor is present in outer quadrants (i.e., at 2, 3 or 4 O’ clock position of left breast or at 8, 9 or 10 O’clock positions of the right breast), dual pedicle technique is preferentially used. In this technique, the inferior pedicle fills up the gap and the NAC is positioned on a superior, superomedial or lateral pedicle. The main aim of the dual pedicle technique is to contour the defect with one pedicle and position the NAC on the other, thereby providing a dual vascular supply.

#### 2.7 Axillary Management

Once these wise pattern incisions are carried through to the chest wall, the lateral dissection is taken into the axilla and the axillary dissection or sentinel node excision is performed through the same incision. No separate incision is taken on the axilla. Care is taken to dissect out the lateral thoracic artery and to ensure that the lateral pillar is well perfused by various perforators. Thereafter, the incisions are closed. Drains are not inserted in the axilla unless an axillary clearance has been performed.

## Results

### 1. Overview of TM Study Cohort: Characteristics of Study Cohort

At our institution, data collection was performed according to the recommendations of the TeaM study protocol. Demographic distribution of study participants and their clinicopathological characteristics are summarized in Figure 2a and Table 2. At our centre 226 TM procedures have been performed on 207 patients with moderate and large breasts with various grades of ptosis during the period of 2012-2019. Amongst these 207, 176 were unilateral breast cancer patients, 12 patients were identified with unilateral benign disease. Among the bilateral cases, 13 were bilateral breast cancer cases, 5 patients presented with one side benign, one side malignant and 1 patient was bilateral benign case.

**Figure 2:**
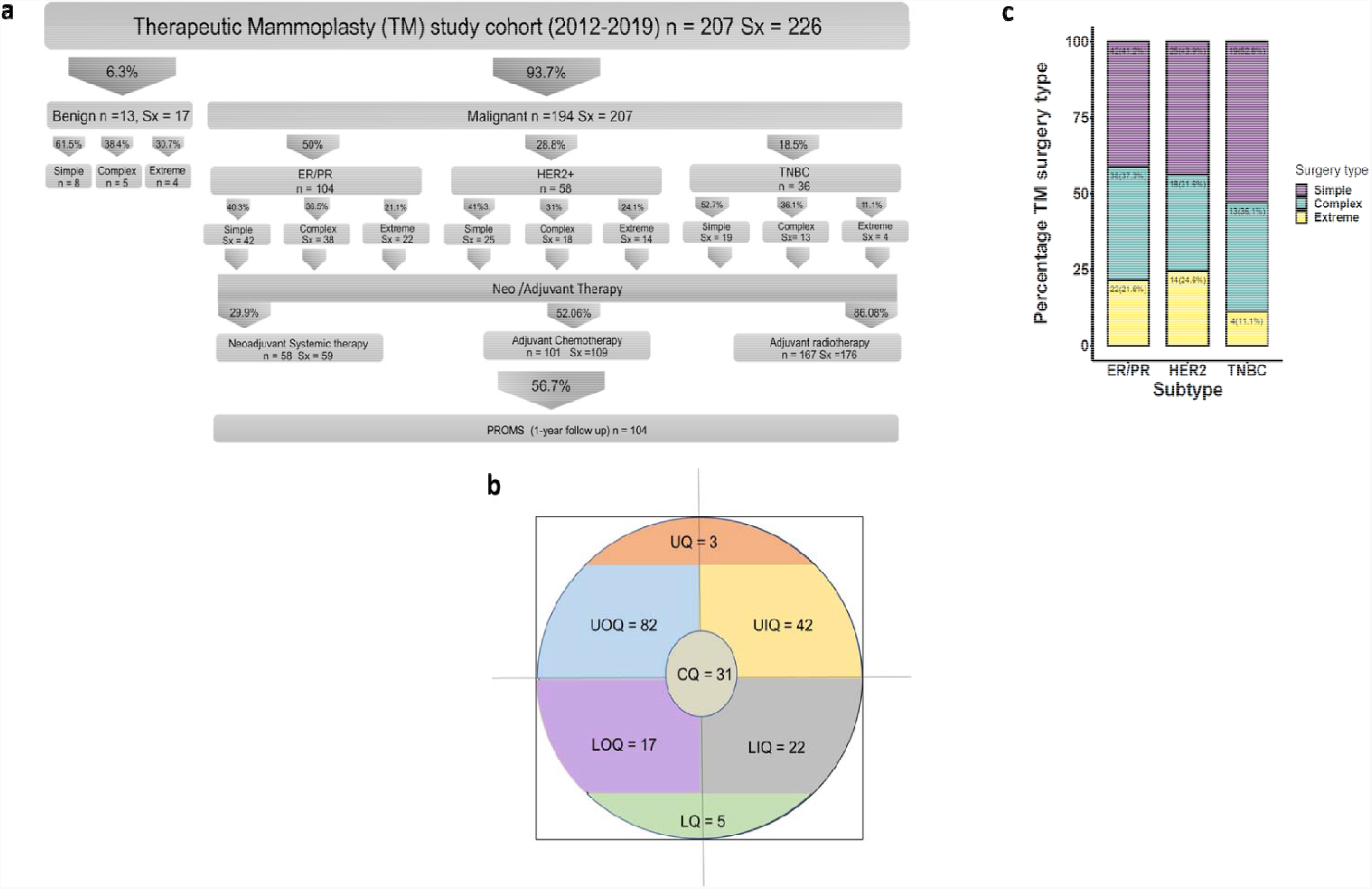
a. Clinicopathological features of the cohort **b**. Quadrant-wise tumor location (CQ – central quadrant, LQ – Lower Quadrant, LIQ – Lower Inner Quadrant, LOQ – Lower Outer Quadrant, UIQ – Upper Inner Quadrant, UOQ – Upper Outer Quadrant, UQ – Upper Quadrant) **c**. Molecular subtype-wise distribution of various TM techniques.

**Table 2:**
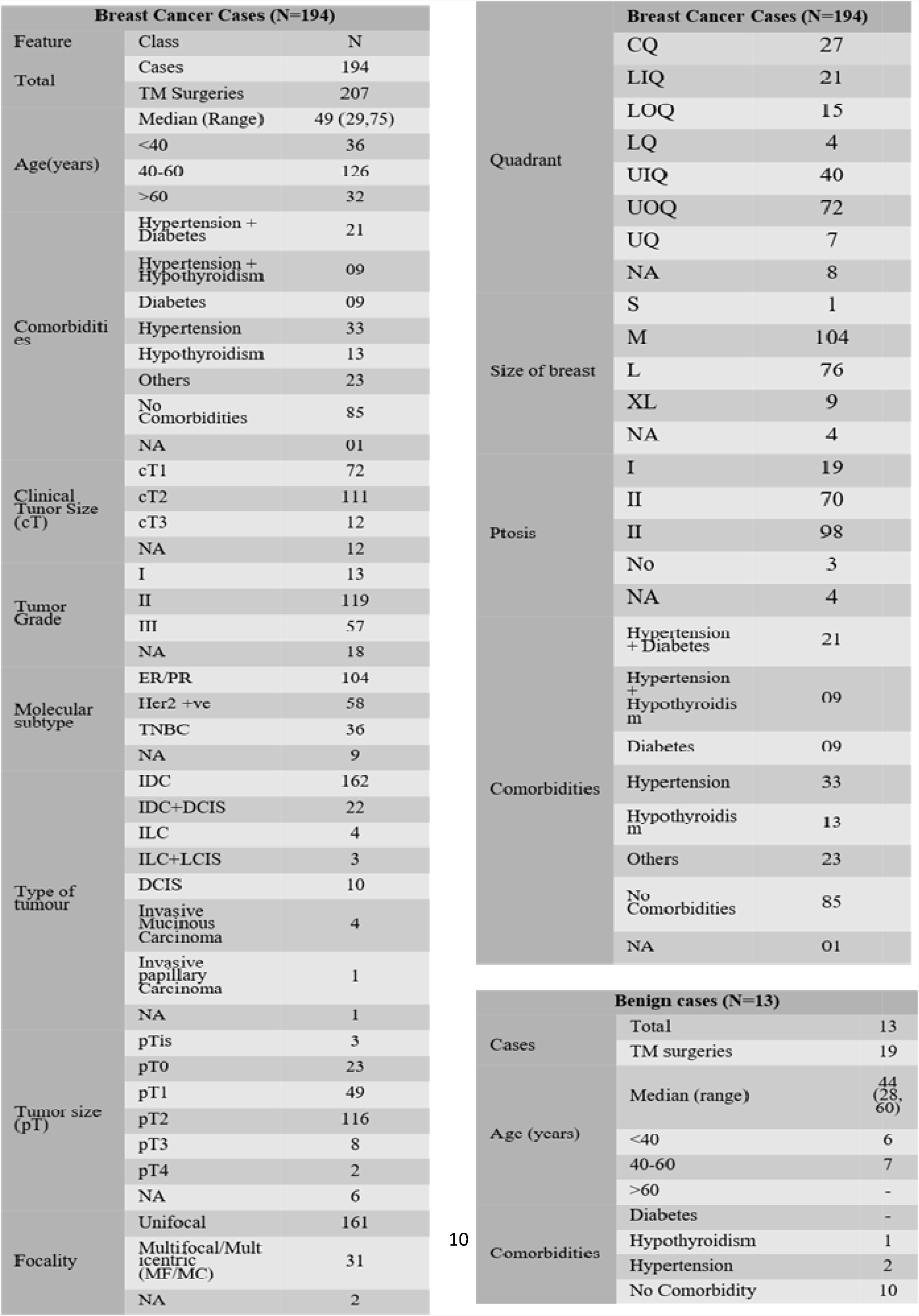
Demographic and Clinico-pathological characteristics of the TM cohort

The median age at diagnosis was 49 years indicating a younger demographic than the overall breast cancer cohort at our centre with 52.7 years as median age from 799 breast cancer cases^23^. As observed in previous reports^24^, a majority of the breast cancer patients (i.e., 109/194 (56.2%) had comorbidities such as obesity and diabetes making them ineligible for mastectomy with immediate reconstruction. Quadrant-wise tumor location is represented in Figure 2b, 61.35% tumors were observed in the upper quadrant. Simple TM accounted for 94 (8 benign) surgeries, while 74 (5 benign) complex and 44 (4 benign) extreme surgeries were performed. Subtype distribution for the different surgeries is shown in Figure 2c. In our study, 127/207 (61.35%) patients had tumors in the upper quadrant, amongst which 82/127 (64.6%) were only in the UOQ.

### 2. Surgical outcomes

#### 2.1 Surgical margins

Clear margins were achieved in all the cancer patients with average margin distance of 12.5 mm. Axillary lymph node dissection was performed in 93 (44.9%) and sentinel node biopsy in 107 (51.6%) of the 207 malignant surgeries.

#### 2.2. Post-Operative Complications

Post-operative complications were classified based on grades as per Clavien-Dindo Classification adapted for breast cancer^25^. A total of 26/194 (13.4%) cases of complications were observed, similar to observations reported in earlier literature^2^ (Figure 3a-b and Supplementary Table 1). All complications were treated conservatively in the outpatient settings. In general, we observe low rate of Grade I/II complications after complex and extreme mammoplasty techniques.

**Figure 3:**
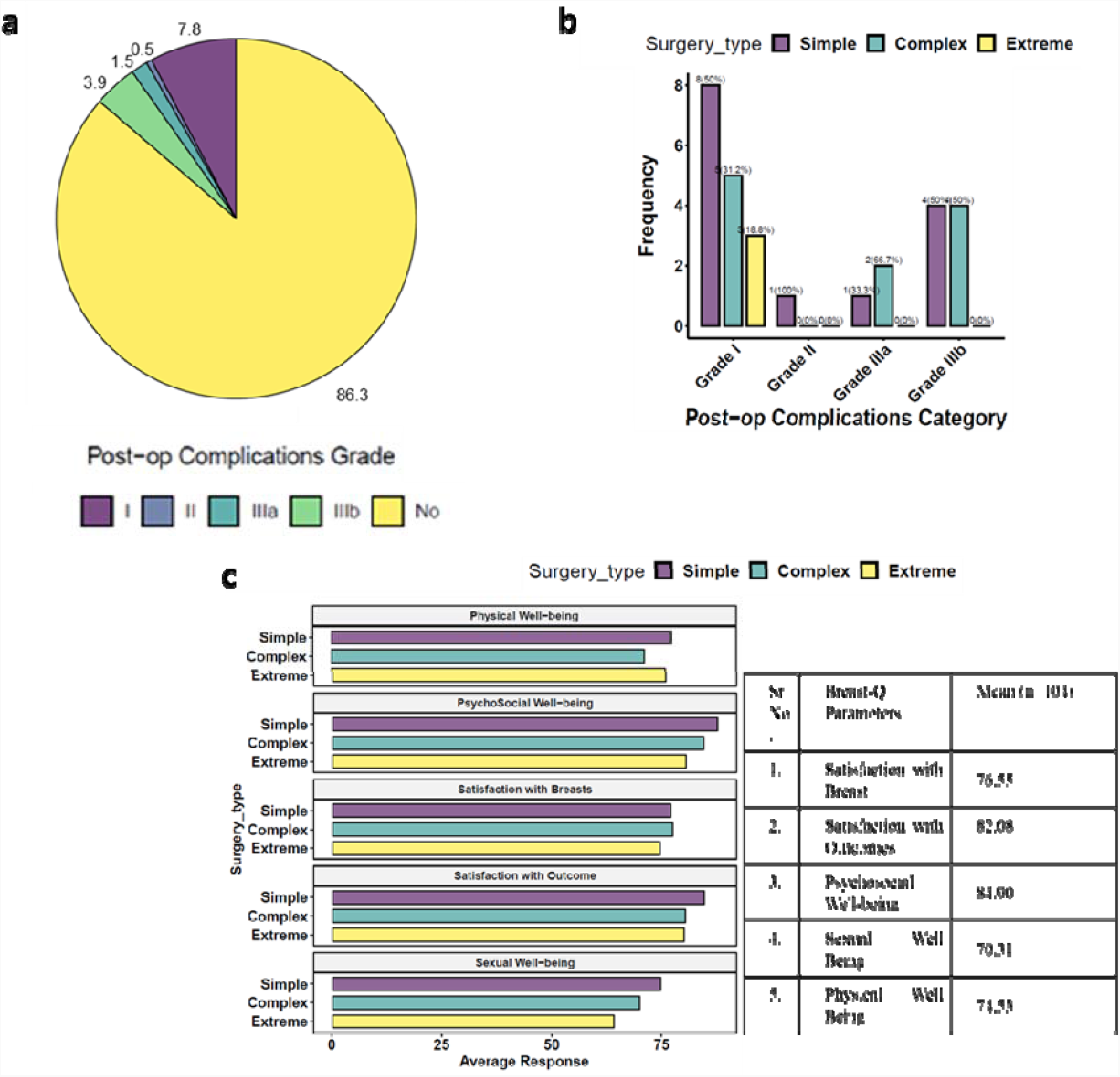
a. Post-op complications observed in the study cohort (in percentage) **b**. Distribution of post-op complications according to TM surgery type **c**. PROMs scores according to TM surgery type represented graphically. Overall PROMs scores presented in the table.

#### 2.3 Patient Reported Outcome Measures (PROMs)

PROMS data was collected from the study participants after a minimum period of 12 months post-surgery using the Breast-Q questionnaires. Out of 207 study participants, 104 (50.2%) responded to the questionnaire. High patient satisfaction scores were observed from our PROMs data as seen in Figure 3c.

#### 2.4 Cosmetic Score Analysis

Out of 207 TM cases for 194 breast cancer patients, cosmetic scores were available for 136 patients based on independent assessment by 3 surgeons. Table 3 shows the observed cosmetic scores.

**Table 3:**
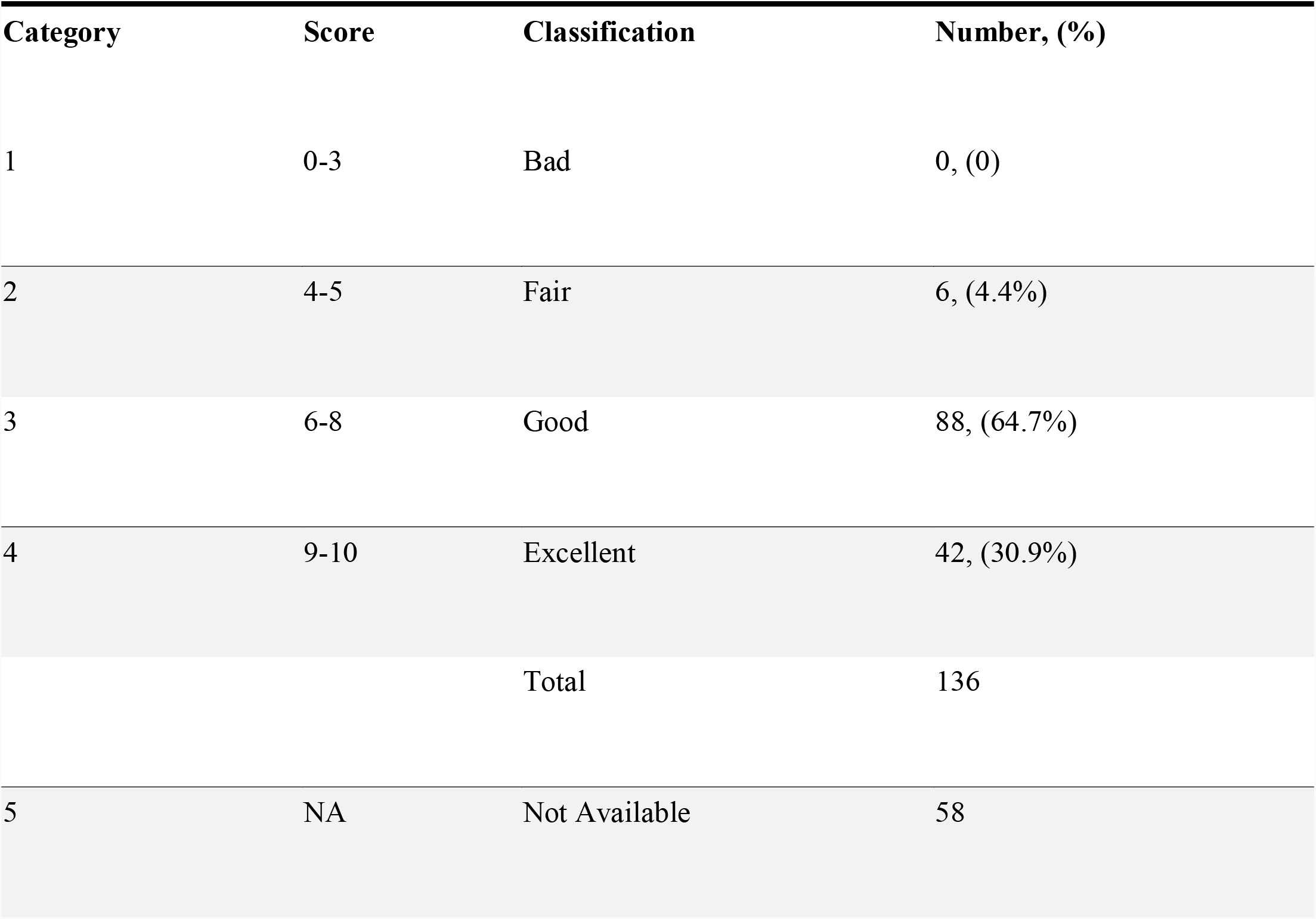
Cosmetic Scores

#### 2.5 Survival Outcomes

The median follow-up for the cohort was 25 months. At median follow-up 3 patients had recurrence with 1 distant and 2 local recurrences.

## Discussion

In this report, we present the first comprehensive, single-institutional study on TM outcomes in breast cancer patients from India based on the recommendations of the TeaM protocol^17^. India brings about its own challenges due to clinical, psychosocial and economic profiles of breast cancer patients being significantly different than western countries. We thus had to adapt our counselling methods for oncoplastic procedures like TM accordingly. Owing to financial and logistical challenges, Indian patients have reduced acceptance of a second operative procedure^26^. Therefore, we perform a single-step TM procedure that involves simultaneous reconstruction of the NAC and contralateral reduction mammoplasty for bilateral symmetrization in which the nipple may undergo resection with a NAC graft. If the patient does not consent for opposite symmetrization, alternative OBS procedures to TM are recommended.

TM-related decision-making algorithms are based on the tumor location and breast size. Most of the cases in our study cohort have been operated using the dual pedicle technique in which NAC was carried on superior pedicle and the inferior pedicle was used to fill the defect caused by excision of the tumor. In patients with smaller breasts or with large excisions, we have frequently used the whole lower segments of the breast (i.e., inferomedial and inferolateral pedicles) so the breast mound is advanced into the defect and NAC is reimplanted onto the pedicle.

Our cohort includes 24 (12.3%) patients who underwent an upfront extreme oncoplasty and 18 (9.2%) patients received NACT followed by extreme TM. In agreement with literature, we recorded a relatively low rate of overall complications. In a post-NACT cohort at our centre, TM was found to be an essential tool to increase the scope of BCS in patients who have large residual tumors (large excision volume). Interestingly, a large majority of the extreme TM performed in this NACT cohort would have qualified for mastectomy at presentation (Koppiker et al., unpublished observations). The potential advantage of TM is wide margins of excision, thus, achieving lower rates of re-excision^27^. Literature reports indicate rates of positive margins range from 0-36% with institutions reporting a 0% margin rate after conducting intra-operative frozen section analysis^28^. In the TeaM protocol, 79% have clear margins^17^. We observed lower rates of margin positivity in our TM cohort which was consistent with previous reports^29,30^. Thus, TM may be a feasible surgical option for breast cancer cases with advanced stages with either MF/MC tumors or LABC at initial presentation^31^.

Re-excision in a case of OBS is challenging due to glandular re-arrangement during mammoplasty and should be considered carefully after discussing within the MDT, if the operating surgeon is confident of identifying the tumor bed and orientation^32^. In our study, only 3(1.5%) patients were reported to have close margins. Re-excision of margins was carried out in one patient, one patient underwent complete mastectomy, and one received an additional boost to the tumour bed.

We also discuss representative cases from each type of surgery. Simple TM typically utilizes a single pedicle and is represented in Case 1 (Supp. Fig. 1). An extended inferior pedicle or a dual pedicle provide optimal outcomes in complex TM procedures (Case 2 and 3; Supp. Fig. 2 and 3). Extreme or split reduction TM is a suitable option for cases with large excisions which were otherwise indicated for mastectomy (Case Study #4; Supp. Fig. 4).

The surgeon’s recommendations for optimum post-operative outcomes are detailed in supplementary information.

In our study, we observed 2 cases of local recurrences (1.03%) and 1 cases of distant recurrence (0.51%) at median follow up of 25 months. Based on these oncological outcomes, we conclude that TM is a oncologically safe procedure even in high-risk patients^33^. Previous reports have indicated that satisfaction with breasts was better in women who underwent OBS than in those who underwent a BCT alone^19^. Overall, we report high levels of satisfaction on PROMs, which is expected^34^, as the aim of TM is to provide an aesthetically pleasing breast. In our study a comparison of the PROMs amongst the types of TMs demonstrates almost equal scores indicating that even complex techniques are well accepted. We also report a higher mean score of 70.3% for sexual wellbeing which may be attributed to better body image and self-esteem arising from the satisfactory outcomes from TM procedure and contralateral reduction mammoplasty. In support of the PROMs, independent assessment by 3 different surgeons indicated that over 90% cases exhibited good-to-excellent cosmetic outcomes

Our encouraging results could be credited to a multitude of factors at our institution such as adequate counselling, shared decision-making, surgical expertise, lower complication rates, personalized hospital services and optimal post-operative care for a long duration.

Our study represents the first detailed report on surgical, oncological and PROMs outcomes after TM surgery in breast cancer patients from India as observed in a single breast oncoplasty unit. We conclude that our TM technique(s) maybe suited for even advanced stage patients with moderate-to-large breasts with mild/severe ptosis. In general, our study observations are compliant with the guidelines of TeaM protocol except few non-compliances such as lack of MRI which has poor uptake in India due to cost barriers.

It may be inaccurate to extrapolate the study findings to the general Indian population due to the large variability in socio-cultural, psychological, and economical ground realities across the country. Similar TM-focused studies from other Indian breast units will be needed in future to corroborate the observations from our study. Indeed, protocols and surgical techniques established here along with PROMs would be a useful framework for replication by other breast surgeons. If the techniques and outcomes of OBS are popularized and the broad indications of TM are clearly defined, it is possible that more eligible breast cancer patients will receive the benefits of this procedure over the routine practice of mastectomy that is currently practiced.

## Data Availability

The study investigators would like to state that all data related to the manuscript will be available for further review on request after consultation with institutional ethics committees

## ABBREVATIONS (arranged alphabetically)

BCS: Breast Conservation Surgery
BCT: Breast Conservation Therapy
DCIS: Ductal Carcinoma In Situ
EO: Extreme Oncoplasty
IDC: Intraductal Carcinoma
MF/MC: Multifocal and/or Multicentric
NAC: Nipple Areolar Complex
OBS: Oncoplastic Breast Surgery
PROMs: Patient Reported Outcome Measures
QoL: Quality of Life
RT: Radiation Therapy
SIB: Simultaneous Integrated Boost
TRM: Therapeutic Reduction Mammoplasty
UOQ: Upper Outer Quadrant

## Acknowledgements

The study authors would like to thank all participants who consented to participate in this study. We acknowledge Bajaj Auto Ltd. for providing support to research activities at Prashanti Cancer Care Mission, Pune. We are grateful to the MAPI Research Trust for permission to use BREAST-Q (http://www.mapitrust.org) and the support from the management and staff of Ruby Hall Clinic, Pune where all surgeries were performed. We would also like to sincerely thank the staff at PCCM for the continued support and efforts.

## Role of the Funding Source

Bajaj Auto Ltd. (GC 0529 and GC 2528) provides funds to PCCM for its research arm, Centre for Translational Cancer Research (CTCR). CTCR is focused on surgical, clinical and translational research activities aimed at addressing problems relevant to breast cancer patients from India.

## Consent

The study was approved by an Independent Ethics Committee. All subjects gave their consent for the use of their personal and medical information including images in the publication of this study.

## Conflict of interest statement

The author declares no competing interests.

## Notes

### Competing Interest Statement

The authors have declared no competing interest.

### Author Declarations

Independent Ethics Committee at Prashanti Cancer care Mission. The study was approved by an Independent Ethics Committee (DCDI/CDSCO registration number: ECR/298/Indt/MH/2018). All subjects gave their consent for the use of their personal and medical information including images in the publication of this study.

