## Supplemental information for "Therapeutic mammoplasty as a viable surgical approach in Breast Cancer Patients from India: A Single Institutional Audit"

**Supplementary Information 1: Methods**

**1. Clinical Management**

Breast cancer diagnosis was based upon clinical examination and radiological evaluation of breast and axilla. Histopathological studies on tru-cut biopsy samples (majority of cases) or vacuum assisted biopsy samples were performed for confirming diagnosis of breast carcinoma. Similarly, ultrasonography and fine needle aspiration cytology was used for investigating axillary lymph node metastasis. Confirmed breast cancer cases underwent breast surgery at a network hospital site. The oncologic management with chemo-radiation protocols was undertaken by a multi-disciplinary clinical team in accordance with the current NCCN guideline^24^.

**2. Assessment of Post-Surgery Complications**

Post-surgery outcomes were assessed by breast oncoplastic surgeons and radiation oncologists, respectively. As per the Clavien Dindo classification, post-surgery complications were classified as ‘major’ when they required surgical intervention and ‘minor’ when they were managed conservatively^23^. We also noted the time between completion of surgery and start of adjuvant therapy to ascertain any delays in adjuvant therapy.

**3. Patient Reported Outcome Measures (PROMs)**

The PROMs were used to evaluate patient satisfaction and QoL after TM procedures. To assess PROMs, a standardized Breast-Q questionnaire was utilized. The Breast-Q module was divided into multiple independent scales. Higher scores indicate greater patient satisfaction and functionality^32^.

**4. Data Analysis and Statistics**

Proportional analysis was undertaken for determining cohort characteristics. Statistical analysis for inter and intra-group comparisons were undertaken using Students t-test (p=0.05).

**Supplementary Information 2: Radiation Therapy**

**RT Methodology**

The breast along with the supraclavicular region (if indicated) was irradiated by 6 MV photon beams using Forward Plan Field- in Field Intensity Modulated Radiation Therapy (F-P FiF IMRT) or Volumetric Modulated Arc Therapy (VMAT). VACLOC immobilization and CT based contouring and planning was done after target delineation after EclipseTM treatment planning system (TPS)(Version 13.5.35) for F-P FiF IMRT plans and Monaco (Version 5.11) TPS for VMAT plans. Tangential fields with sub-fields were used for radiotherapy planning. Linac, Elekta Medical SystemTM, UK with 80 leaves Multi Leaf Collimator (MLCi)was used. RT plan was accepted if atleast 95% of prescribed dose covers the 100% of planning target volume (PTV). Hot spot in PTV was accepted up to 110%of the prescribed dose. Tumour bed boost, wherever indicated, was performed using either an electron portal or Simultaneous Integrated Boost (SIB) technique with standard dose fractionation schedules.

**RT Results**

Of the 194 breast cancer patients included in our study cohort, 169 patients underwent RT as clinically indicated. Among the 194 patients, 42 did not have any adverse reactions to the radiation therapy, while 81 developed Grade I reactions, 30 showed Grade II reactions and 5 Grade III reactions, for 11 patients’ information post radiation therapy was not available. The RT regimen for various types of TM procedures was thus considered effectively safe.

**RT Discussion:**

In our opinion, in majority TM cases, margins around the tumor bed do not shift significantly due to following reasons:

- During TM, adequate care is taken to check whether tumor bed is well delineated with markings by Liga clips, as soon as tumor is removed.
- In some cases, the margins may get advanced into the tumor cavity to form the bed of the tumor cavity (such as in an extended inferior pedicle). Herein, for dealing with the tumor in the superior quadrant, the lower margin (which is the highest point of the extended pedicle) shifts into the tumor cavity, where exactly the boost is required.
- In simple mammoplasty (or tumor in lower quadrant or superior quadrant), in which the tumor is in a tissue segment within the specimen, it is likely that some of the margins may shift into the tumor cavity but not shift away from it.
- If the tumor is lying outside (i.e., in outer quadrant or supero-medial quadrant) and if the excision is large, central mound advancement can be performed to fill up these cavities. In this situation, even if the infero-medial margin may shift, being a supero-medial margin, but it will not go outside the tumor cavity.
- For cavity on the outer side, if a dual pedicle technique is applied, even then inferior pedicle will be used only to fill in the gap.

Guidelines for optimal RT planning after TM are unclear and further methodical investigations are needed. Indeed, results are eagerly awaited from the MIAMI trial which is the first randomized trial design to address the clinical safety of TM associated with the excision of each cancer and the possibility of performing up to two tumor bed(s) boost(s) radiotherapy (ClinicalTrials.gov Identifier: NCT03514654).

In our study, the mean duration from TM to start of adjuvant treatment was 50 days without any delay. This observation is consistent with several studies that indicate OPBS does not result in delay in adjuvant treatment^3^. The optimal duration between OPBS and RT has not been established. In our practice, we prefer commencing RT within 6 months of treatment in which adjuvant chemotherapy has been administered. If no ACT is required, we start RT within 5-6 weeks.

**Supplementary Information 3: Surgeon’s Recommendations**

- Careful marking placement, so that the closures are not tight.
- The tumour excision should be maximum through one limb of the incision and axilla should be accessed through the same incision by identifying the lateral border of the Pectoralis major and minor.
- The Supero-lateral area and the lateral pillar should be carefully mobilized to prevent devascularization from the lower lateral segment.
- SLNB should be performed through the same incision using ICG or nuclear dye and if the status is positive, axillary dissection should be carried out via the same incision.
- All the tumour margins should be analysed on frozen sections and by a specimen mammogram. The breast restoration should be delayed until the results on frozen section are negative. The contralateral reduction should be performed while frozen sections analysis is ongoing.
- The interruptive sutures should be used at the ‘T’ junction instead of continuous sutures to minimize necrosis.
