## Supplemental table for "Therapeutic mammoplasty as a viable surgical approach in Breast Cancer Patients from India: A Single Institutional Audit"

Table 1: Post-op Complications in the cohort as per the Clavien Dindo Classification

| **Characteristics** | **Complications, Number % (n=194)** | | | |
| --- | --- | --- | --- | --- |
| **Grades** | **Total**  **(26)** | **Simple TM**  **(13)** | **Complex TM (11)** | **Extreme Oncoplasty (2)** |
| **Grade 1 (**seroma/hematoma not requiring drainage, minor skin necrosis, fat necrosis, delayed wound healing) | 17 (8.7) | 7 (3.6) | 9 (4.6) | 1 (0.5) |
| **Grade 2**  (Wound infection) | 6 (3) | 4 (2.06) | 1 (0.5) | 1 (0.5) |
| **Grade 3a (**seroma/hematoma which were drained under USG guidance, lymphoedema, nipple necrosis, skin necrosis undergoing debridement) | 2 (1.03) | 1 (0.5) | 1 (0.5) | 0 |
| **Grade 3b** (seroma/hematoma which were drained under general anesthesia – major skin necrosis, wound infection requiring debridement, bleeding) | 1 (0.5) | 1 (0.5) | 0 | 0 |
| **Total** | 26 (13.4) | 17 (8.7) | 8 (4.1) | 2 (1.03) |
